# Perceptions of young people in the Democratic Republic of Congo on the factors influencing their intention to use mobile health applications for comprehensive sex education: A descriptive qualitative study

**DOI:** 10.64898/2026.03.16.26348571

**Authors:** François Kajiramugabi Maneraguha, José Côté, Anne Bourbonnais, Caroline Arbour, Marie Hatem

**Affiliations:** Faculty of Nursing, Université de Montréal, Montreal, Quebec, Canada; Chaire de recherche innovante en santé durable et sciences infirmières en Afrique, Bukavu, South Kivu, Democratic Republic of the Congo; Institut supérieur des techniques médicales de Bukavu, Bukavu, South Kivu, Democratic Republic of the Congo; Research Centre of the Université de Montréal Hospital Centre, Montreal, Quebec, Canada; Canada Research Chair in Care for Older People, Montreal, Quebec, Canada; Research Chair in Nursing Care for Older People and their Families, Montreal, Quebec, Canada; Research Centre of the Institut universitaire de gériatrie de Montréal, Montreal, Quebec, Canada; School of Public Health, Université de Montréal, Montreal, Quebec, Canada; International Center for Advanced Research and Training, Fondation Panzi, Bukavu, South Kivu, Democratic Republic of the Congo

**Keywords:** Intention to use, Mobile health applications, Comprehensive sex education, Young people, Bukavu

## Abstract

**Background:** Young people’s access to quality comprehensive sex education (CSE) is crucial to promoting their health and well-being. This access remains limited in the Democratic Republic of Congo, despite increasing use by young people of smartphones and social media, which does not lead to structured access to digital CSE. In this context, mobile health applications represent a promising solution, as they complement interpersonal CSE by providing confidential, accessible and culturally appropriate learning. However, few studies have explored young people’s perceptions of the factors influencing their intention to use such an application. The purpose of this study is to fill this gap with young people interviewed in the city of Bukavu.

**Methods:** A descriptive qualitative study was conducted with 21 students recruited by purposive sampling from eight public secondary schools in Bukavu. Development of the interview guide was informed by the Unified Theory of Acceptance and Use of Technology. Semi-structured, face-to-face individual interviews were conducted until thematic saturation was reached, with audio recordings analyzed using Braun and Clarke’s inductive thematic approach, supported by MAXQDA Pro 24.8.0 software.

**Results:** Six themes and twenty interrelated subthemes emerged: (i) intention to use based on personal need; (ii) structural, economic and psychosocial barriers; (iii) adaptive financial strategies for accessing digital CSE; (iv) the positive influences of families and peers; (v) perceived educational usefulness of the applications; and (vi) perceived conditions for digital engagement. The young people expressed a need for reliable information. They develop ingenious access strategies to offset the cost of mobile data, smartphones and repeated power outages. Their intention is also based on financial and technical support from friends and family. They perceive such applications as useful, especially in the context of a humanitarian crisis. Their perceived engagement depends on ease of use, familiarity with digital technologies, information reliability and offline access.

**Conclusions:** This study enriches the field of digital health by showing that the intention to use CSE applications is based on both favourable perceptions and facilitating and adaptive conditions, including perceived educational benefits, innovative access strategies and a mobilizing social environment. It highlights the need to co-design, with young people, solutions tailored to their needs to guide the development of realistic digital interventions that are sensitive to the local context.

**Author Summary:** In this study, we sought to understand what motivates—or discourages—young people in Bukavu, in the Democratic Republic of Congo, from using mobile health applications to learn about the body, relationships, and infection prevention, adolescent pregnancy, and violence. We met with 21 students from eight public secondary schools and asked them, in individual interviews, what they expected from such tools.

First and foremost, the young people expressed a strong need for reliable information on sexual health, provided discretely, especially since these topics are rarely discussed at school or in the home. They also described very real obstacles: the cost of an Internet connection, smartphone availability, repeated power outages, fear of coming across inappropriate content, and concerns about confidentiality. They reported some ingenious strategies for overcoming these obstacles, such as saving money, taking small paid jobs, using their neighbours’ Wi-Fi and connecting to an external battery.

Our findings demonstrate that a truly useful mobile health application must be simple, accessible offline, inexpensive, and tailored to local conditions. They also suggest that the best way to achieve this is to design such solutions in collaboration with young people, so that the solutions meet their needs and reflect the reality of their daily lives.

## 1. INTRODUCTION

Comprehensive sex education (CSE) is a key public health strategy for improving the sexual and reproductive health and well-being of young people and informing them about their rights in this area (1). It makes a direct contribution to the achievement of several Sustainable Development Goals, including health, quality education, gender equality, poverty reduction and economic growth (2). CSE is evidence-based. It strengthens young people’s knowledge, attitudes and skills around eight essential components: relationships; values, rights, culture and sexuality; understanding gender; violence and safety; health and well-being skills; the human body and development; sexuality; sexual and reproductive health (SRH); and sexual behaviour (2). Recent systematic reviews show its effectiveness in promoting condom use and responsible sexual behaviour, reducing sexual violence, preventing adolescent pregnancy, and supporting reproductive justice (3–5).

Nevertheless, the implementation of CSE in French-speaking sub-Saharan Africa (FSSA) remains limited by sociocultural and structural barriers (6,7). In the Democratic Republic of Congo (DRC), only 17% of young people had access to CSE in 2021 (8). This low level of access can be traced to sociocultural norms that limit discussions about sexuality, unequal gender relations, and the limited availability of educational and sexual health services tailored to young people’s needs (9–11). This deficit perpetuates unequal norms, to the detriment of young people’s health, well-being and sexual rights. Among Congolese girls, this translates into a very low level of preparedness for the onset of menstruation (12) as well as a high proportion of unwanted pregnancies (estimated at 80%), unsafe abortions (49%) and early marriages (15). It is accompanied by alarming rates of maternal mortality (15), sexual violence (16), low levels of knowledge about contraception (11) and a low prevalence of contraceptive use (28%) (17,18). Among boys, the gaps are related to knowledge and attitudes toward condoms and positive masculinity (19,20).

In this context, digital environments offer young people with new opportunities to access CSE. Living in a highly digitized world, they use the Internet and mobile phones on a daily basis as vehicles for socialization and empowerment (21,22). Digital CSE is therefore emerging as a promising and innovative response (2) that complements school and family-based approaches, especially when these are ineffective or inaccessible (23). It relies on digital interventions, such as mobile health applications, designed to work on smartphones, tablets and other mobile devices (24). Driven by mobile health applications, digital CSE is now a cornerstone of the modernization of health services for young people. It helps to overcome taboos and ensure confidentiality and can reach young people in their everyday environments(23). A recent mapping of evidence shows that, in today’s digital landscape, mobile health applications embody innovation, accessibility and relevance in meeting the CSE needs of African youth (25). This threefold nature is based on their ability to provide interactive, engaging and personalized content, thereby reinforcing young people’s intentions to use them. As such, two overview studies (26,27), and nine systematic reviews—quantitative, qualitative, or mixed— (25,28–35), have synthesized more than 400 studies conducted between 1987 and 2023 in low, middle, and high-income countries. This synthesis has highlighted the ability of digital CSE to strengthen skills and behaviours that promote the health and well-being of young people. However, despite this strong evidence base, uncertainties remain with respect to their contextual relevance. An analysis of these 11 reviews (25–35) revealed two key findings. First, the majority of the studies were conducted in English-speaking or high-income contexts, limiting their transferability to low-income French-speaking African contexts. In sub-Saharan Africa, the research remains heavily concentrated in English-speaking countries such as South Africa, Nigeria, Kenya, Ethiopia, Liberia, Ghana, Tanzania, Rwanda, Zambia and Malawi. In contrast, one study was identified in each of the following French-speaking countries: Senegal, Cameroon, Mali, and RDC (26,27,36). On the other hand, young people’s intentions to use mobile health applications for their CSE remain understudied: the available studies focus more on experience of use rather than intention to use. However, Kim and Park (37) argue that intention to use is a key factor in the acceptance of health technologies: it determines actual use, user engagement and the quality of the data generated, so it influences the effectiveness and efficiency of digital interventions. The importance of analyzing this intention in the lead-up to use is also highlighted by Perski and Short (38), who point out that the acceptability of digital interventions, such as mobile health applications, is not limited to the user experience: it encompasses both affective attitudes toward the tool and the intention to use it, which precedes actual use. The available qualitative reviews (33–35,39) confirm this gap: the few existing studies focus mainly on other forms of digital interventions, such as web platforms, SMS services and online educational programs, but do not explore young people’s intention to use mobile health applications for their CSE. Thus, no study has examined how young people structure their motivations, perceived barriers, strategies for accessing mobile data, or the role played by friends and family in this intention, even though these dimensions are major contextual factors in FSSA. This finding highlights a significant gap between the proven effectiveness of digital CSE and our lack of knowledge about its potential acceptance by young people (40,41). Contextual adaptation is essential for the DRC, where connectivity is growing rapidly, characterized by a 52% mobile penetration rate and 27% mobile Internet access rate (42), rapid growth in Internet users (+122% from 2019 to 2020), and the marked popularity of WhatsApp (43). Among young people in the Congolese provinces of North and South Kivu, social media use is intensifying in a context of humanitarian crisis (44). This familiarity, combined with persistent barriers such as connection costs, poor digital literacy and irregular energy availability, suggests that usage intentions are shaped by technical, social and economic factors that have still not been well documented. Given this context, the development of health informatics and digital technology in the DRC (45,46) represents a strategic window of opportunity (47) for strengthening the provision of digital CSE and promoting the sexual health of Congolese youth. This group, which represents more than 65% of the population (48), plays a key role in realizing the demographic dividend (49). Paradoxically, their intention to use mobile health applications for their CSE has not been sufficiently described: the relatively few available studies are on chatbot messaging (43) and voice broadcasting (50). Therefore, understanding how Congolese youth navigate technical, financial and relational constraints, and how they perceive the educational relevance of these applications, represents an essential prerequisite for developing appropriate interventions. This dual gap—low Francophone representation and a paucity of studies on intention to use—limits the ability to design mobile health applications that are truly rooted in local realities. This study aims to fill this gap by describing the perceptions of Congolese youth regarding the factors influencing their intention to use mobile health applications for their CSE.

## 2. METHOD

### 2.1. Design

To achieve the objective of this study, a qualitative descriptive approach was adopted (51) involving an in-depth exploration of the expressed perceptions of young people and based on the Unified Theory of Acceptance and Use of Technology (UTAUT) (52). This theory posits that the use of a technology is determined by behavioural intention, which is itself influenced by four factors: performance expectancy (perceived usefulness), effort expectancy (perceived ease of use), social influence (expected support from peers) and facilitating conditions (availability of resources); as well as by three moderators: age, gender and experience. In this study, UTAUT was used to structure the development of the interview guide, not as a coding grid but as a flexible framework guiding the formulation of questions, thus preserving the spontaneity and richness of the participants’ comments (53). The parts of the study are reported according to the COREQ (Consolidated Criteria for Reporting Qualitative Research) grid (54) (Appendix 1).

### 2.2. Setting and participants

The study was conducted in eight public secondary schools in the city of Bukavu, South Kivu province. These schools are located in three municipalities (Bagira, Ibanda and Kadutu) and four educational zones (Bukavu I to IV), including state-funded or religious schools and non-state-funded schools. This distribution reflects the diverse local educational landscape.

The sample consisted of students aged 15 to 24 years. It includes girls and boys in different classes, from the first to the fourth year of secondary education in the DRC. The target age group (15-24 years) was based on the definition of the concept of “youth” recommended by United Nations Resolution 36/28 of 1981 (55) and adopted by UNESCO, UNICEF and UNFPA (56). This choice also takes into account the extended length of secondary education in the DRC and the high dropout rates, which often lead to the presence in secondary school of students over the age of 21 years (57). Participants were selected by purposive sampling, based on gender, place of residence and school attended. School administrators facilitated the initial contacts with participants. Interested students were approached individually by the lead author (FKM). During this presentation, the students were informed of the principal investigator’s role, research objectives, and university affiliation to promote transparency. A total of 21 participants were included (Table 1). None of the pupils approached refused to participate or dropped out during the data collection period.

**Table 1:** Distribution of participants according to school characteristics.

| Gender of participants | Type of School |  |  |  |  |  |  |  |
| --- | --- | --- | --- | --- | --- | --- | --- | --- |
|  | Catholic state-subsidized |  |  | Protestant state-subsidized |  | Non-subsidized |  |  |
|  | Collège Alfajiri | Institut Avenir | Institut Kasali | Institut Bangu | Institut Imani Panzi | Institut de Kadutu | Athénée d'Ibanda | Institut de Bagira |
| Girls | 2 | 1 | 1 | 2 | 1 | 2 | 2 | 2 |
| Boys | 2 | 0 | 2 | 1 | 0 | 1 | 1 | 1 |
| Total | 4 | 1 | 3 | 3 | 1 | 3 | 3 | 3 |

### 2.3. Data collection

The data were collected in the fall of 2024. The interviews were conducted in person by FKM, with support from a research assistant for a greater climate of trust, given the sensitivity of the subject. The interviews were held in classrooms provided by the school administrations, ensuring confidentiality and comfort for the participants. A semi-structured interview guide (Appendix 2) was developed. It included open-ended questions inviting participants to express their perceptions of the factors likely to influence their intention to use mobile health applications for CSE. This guide was validated by the scientific committee and then, to ensure its clarity and relevance, it was pre-tested with six students who were left out of the final sample. The interviews were conducted in a conversational manner, encouraging participants to express themselves freely. Follow-up questions were used as needed to clarify or enrich the responses. They were recorded, with the participants’ consent, using a dictaphone to ensure the accuracy and completeness of the transcriptions. Field notes were taken during and after the interviews to record contextual and non-verbal details. These were then incorporated into the analytical process to enrich interpretation of the verbatim transcripts. The interviews varied in length from 30 to 60 minutes. o repeat interviews were conducted. At around the 18th interview, the discussions were not yielding any new themes, indicating that data saturation had been reached. This observation was confirmed at the end of the 21 interviews. To strengthen the study’s credibility, the principal investigator maintained his reflexivity throughout the process, recording his impressions in a logbook.

### 2.4. Data analysis

The coding was carried out by FKM using an iterative process of continuous rereading and comparison. The codes, generated inductively from significant segments of the transcripts, were then organized into subthemes and main themes. The six-step thematic analysis of the data according to Braun and Clarke (58) was combined with descriptive coding (59) as follows: (1) familiarization with the data (listening to the recordings, full transcription and repeated rereading of the verbatim transcripts); (2) initial coding by assigning a concise word or expression to each segment of meaning that would reflect the segment’s main content without in-depth interpretation; (3) a search for themes achieved by grouping related codes into thematic categories; (4) reviewing and refining the themes, creating sub-codes where necessary to reflect the full range of perceptions; (5) clearly defining and naming each theme and subtheme, supported by illustrative quotes from the verbatim transcripts; (6) final drafting of the results in narrative form, incorporating the most significant quotes to illustrate the dynamics specific to the intention to use mobile health applications for CSE. Coding was performed using MAXQDA Analytics Pro 24.8.0 software to organize and manage the excerpts and thematic categories. The transcripts were not returned to the participants, as the school setting did not allow for individual follow-up after the interviews.

### 2.5. Scientific criteria

The rigour of this study was ensured by applying Lincoln and Guba’s four criteria (60): credibility, transferability, reliability and confirmability. Credibility refers to confidence in the accuracy of the results. It was based on triangulating data sources from the eight schools, achieving thematic saturation and keeping a reflexive journal. Transferability refers to the possible application of the results obtained to other, similar contexts. It was facilitated by a detailed description of the study context. Reliability, which corresponds to the stability of the data and the consistency of the results, was ensured by transparently describing the data collection and analysis procedures, supported by the use of MAXQDA software to archive analytical decisions. Confirmability depends on fidelity to the data. It was ensured by the traceability of the analyses, by using verbatim quotes that illustrate the final themes, and by documenting the analytical choices in an audit trail. It was also reinforced through a process of continuous reflexivity by the principal investigator and regular exchanges with the co-researchers on the consistency of interpretations and the relevance of the thematic groups.

### 2.6. Ethical considerations

The study was approved by the Science and Health Research Ethics Committee at Université de Montréal (CERSES 2024-6039) and the DRC’s National Health Ethics Committee (CNES 001/DPSK/222PM/2024). Before participating, the students were informed of the study’s objectives and procedures, its voluntary nature, and their right to withdraw at any time without harm to their education. Free and informed consent was obtained from participants aged 18 and over through a written form. In accordance with Congolese law, for participants aged 15 to 17, consent was obtained from the school principals, who acted as legal guardians. The students’ consent was then sought and obtained using an information form that they signed before any data was collected. In presenting the results, the anonymity of participants was ensured by assigning an alphanumeric code combining the letter P, a serial number and the letter *F* or *M* to indicate gender. This distinction was relevant because perceptions of digital CSE differ by gender, and this must be considered due to the practical implications of this research.

## 3. RESULTS

### 3.1. Sociodemographic, educational and technological profile of participants

The sample consisted of participants with varied sociodemographic, educational and technological profiles, reflecting the diversity of schools in Bukavu. Table 2 presents their general characteristics.

**Table 2:** Participant profile. (N=21)

| Characteristic | Results |
| --- | --- |
| Age in years (mean ± standard deviation) | 17 years (± 1.44), range: 16 to 20 years |
| Gender | Girls: 13 (62%); Boys: 8 (38%) |
| Communes of origin | Ibanda: 9 (43%); Kadutu: 6 (29%); Bagira: 6 (29%) |
| Residential neighbourhoods | Urban: 8 (38%); Suburban: 13 (62%) |
| Training options | Science, Education, Social Sciences, Nutrition |
| Smartphone ownership | Yes: 18 (86%); No: 3 (14%, but experienced in its use) |
| Phone brands owned | Tecno, Infinix, Pixel, LG, Oppo |

### 3.2. Participants’ perceptions of factors influencing their intention to use mobile health applications for their CSE

#### 3.2.1. Synopsis of themes and subthemes

Six main themes and twenty interrelated subthemes emerged from the thematic analysis. They are presented in Table 3.

**Table 3:**
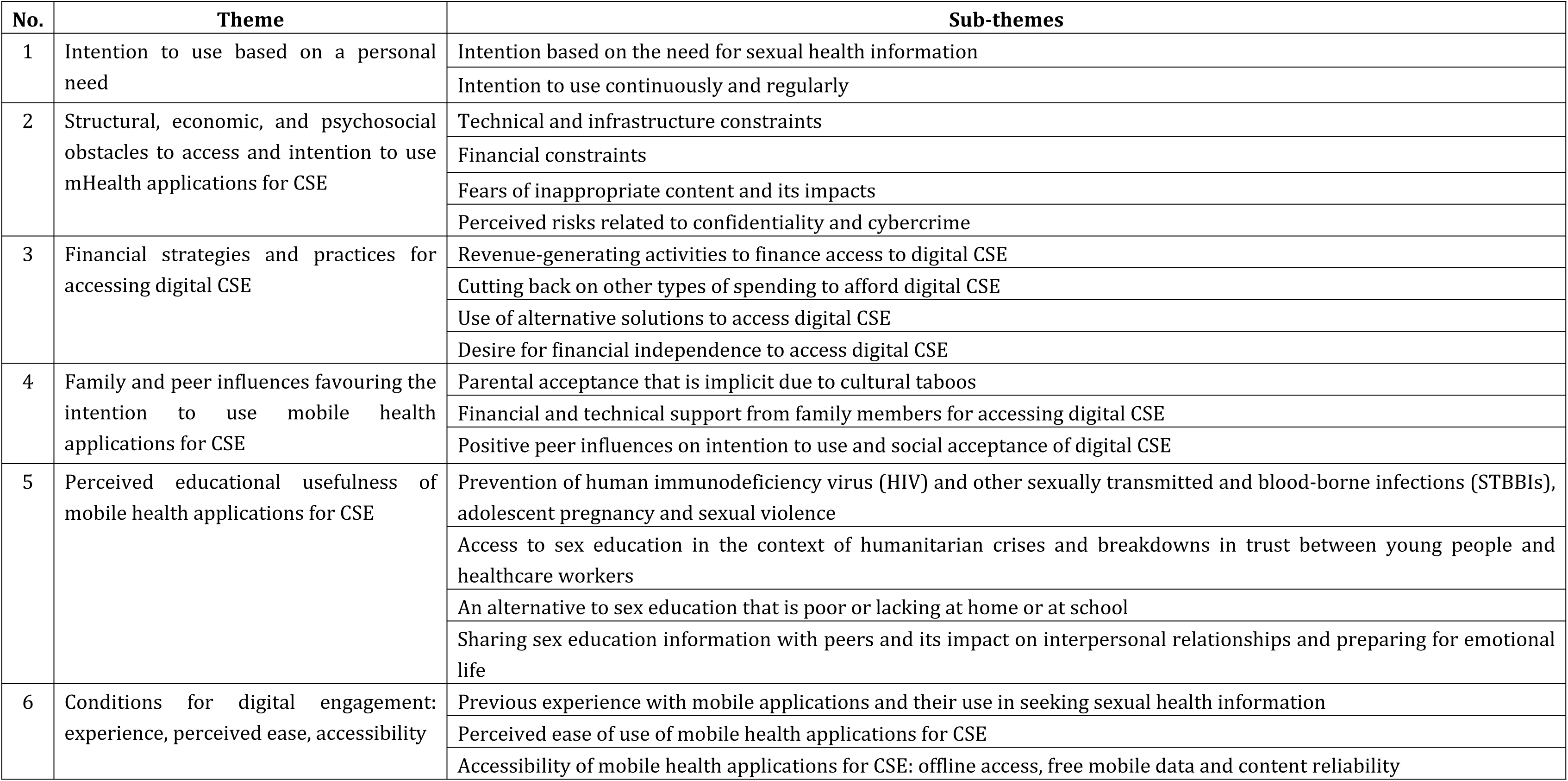
Themes and subthemes identified on factors influencing the intentions of young people in the DRC to use mobile health applications for their CSE.

| No. | Theme | Sub-themes |
| --- | --- | --- |
| 1 | Intention to use based on a personal need | Intention based on the need for sexual health information |
|  |  | Intention to use continuously and regularly |
| 2 | Structural, economic, and psychosocial obstacles to access and intention to use mHealth applications for CSE | Technical and infrastructure constraints |
|  |  | Financial constraints |
|  |  | Fears of inappropriate content and its impacts |
|  |  | Perceived risks related to confidentiality and cybercrime |
| 3 | Financial strategies and practices for accessing digital CSE | Revenue-generating activities to finance access to digital CSE |
|  |  | Cutting back on other types of spending to afford digital CSE |
|  |  | Use of alternative solutions to access digital CSE |
|  |  | Desire for financial independence to access digital CSE |
| 4 | Family and peer influences favouring the intention to use mobile health applications for CSE | Parental acceptance that is implicit due to cultural taboos |
|  |  | Financial and technical support from family members for accessing digital CSE |
|  |  | Positive peer influences on intention to use and social acceptance of digital CSE |
| 5 | Perceived educational usefulness of mobile health applications for CSE | Prevention of human immunodeficiency virus (HIV) and other sexually transmitted and blood-borne infections (STBBIs), adolescent pregnancy and sexual violence |
|  |  | Access to sex education in the context of humanitarian crises and breakdowns in trust between young people and healthcare workers |
|  |  | An alternative to sex education that is poor or lacking at home or at school |
|  |  | Sharing sex education information with peers and its impact on interpersonal relationships and preparing for emotional life |
| 6 | Conditions for digital engagement: experience, perceived ease, accessibility | Previous experience with mobile applications and their use in seeking sexual health information |
|  |  | Perceived ease of use of mobile health applications for CSE |
|  |  | Accessibility of mobile health applications for CSE: offline access, free mobile data and content reliability |

#### 3.2.2. Descriptions of themes and subthemes illustrated by verbatim quotes

##### Theme 1: Intention to use based on a personal need

This theme explores the motivations underlying the use by Congolese youth of mobile health applications for CSE. Two subthemes structure these perceptions.

###### 1.1. Intention based on the need for sexual health information

The intention to use mobile health applications for CSE is based on a personal need for sexual health information, which is perceived as essential to understand how the body works and improve one’s knowledge. As P11F explained: *“When I use the mobile application, I learn things I didn’t know about my menstrual cycle and genital hygiene, and that has helped me improve my knowledge.“* This intention is also described as a proactive approach to meeting personal needs. P2M stated: *“Yes, I’d continue to use them, because I’m looking to meet my own sexual health needs. My goal is to obtain the information I need. I see it as a personal need.“* Other participants mentioned seeking information for prevention. P3F said: *“I’m interested in my physical health, such as sexual intercourse. A mobile health app could help me learn how to protect myself during sex.“* For P10F, this intention is also rooted in social and family expectations: *“I’d use them so that I’ll know how to behave as their only daughter, to not cause [my parents] any shame in my life.“* Lastly, it is expressed as a personal responsibility and a challenge for the future. P13M said: *“I’m going to use them. Yeah, for me, my sex education and sexual health are priorities; my future matters, it’s my business. The information I find there is important for [understanding how] my body works and for managing intimate relations with my future girlfriend.”* According to P20F, this intention is directly linked to physical experiences and prevention: *“Since I’ve already begun to have periods [menstruation], I’m interested in these mobile apps because I can learn how to avoid pregnancy with my boyfriend and how to protect myself against HIV/AIDS.”*

###### 1.2. Intention to use continuously and regularly

For some young people, the intention to use mobile health applications for CSE is part of a plan for continuous and regular use, fuelled by the content they provide and its perceived usefulness. P1M expressed a lasting commitment to these applications: *“I’d recommend giving these mobile health apps to all students my age at our school because they’ve helped me a lot. I don’t know if I’ll ever stop using them.“* This intention to continue using them is also associated with an interest in information that is deemed relevant to sexual health and the body. P3F explained: *“Yes, I’m very interested in mobile health apps. The information about my sexual health and body encourages me to use them all the time. The content that would encourage me even more to use them all the time is when there’s useful information on young people’s sexual health.“* For P4F, this intention was part of a dynamic of communication and sharing around topics she considered important: “*The topics that would encourage me to keep using them are those that allow me to stay in touch with my family and friends to talk about, above all, a variety of interesting things, including genital health and, for girls, how to avoid getting pregnant.“*

These two subthemes therefore show that the intention to use mobile health applications for CSE is based on a personal quest for knowledge about sexual health and is part of a long-term commitment that was widely shared by the participants. There were nevertheless some gender differences: the girls placed more emphasis on physical experiences, pregnancy prevention and social expectations, while the boys were more focused on access to information and long-term use.

##### Theme 2: Structural, economic and psychosocial obstacles to access and intention to use mobile health applications for CSE

Despite their stated intention to use digital CSE, Congolese youth describe obstacles that could hinder this. Presented in four sub-themes, these perceived obstacles relate to technical and infrastructure, financial and psychosocial constraints.

###### 2.1. Technical and infrastructure constraints

The obstacles identified by participants include limited access to electricity, network outages and unstable Internet connections. P3F felt that an unreliable Internet connection could limit her access to content and application updates: *“I’d need a good Internet connection for regular updates to this mobile sex education app.“* P4F pointed out that when the connection is unstable, access to information becomes impossible: *“If my Internet connection is poor, I won’t be able to use these mobile apps to access sexual health information.*” Lastly, P12M mentioned frequent power outages as an obstacle to recharging telephones and using applications on a regular basis: *“I often have problems charging my phone. The power from the National Electricity Company regularly goes out, so we run out of power for our phones.“*

###### 2.2. Financial constraints

The young people mentioned financial constraints related to the high cost of mobile data plans (*megabytes*) and of smartphones, perceived as barriers that may limit their intention to use CSE mobile health applications. P2M said: *“The hardest part is finding megabytes. Besides, I don’t have an Android phone, and I may not have the means to buy the megabytes I need, either, to be able to use these mobile health apps.“* P5F highlighted the recurring nature of this constraint: *“I buy megabytes every week, sometimes two or three times, but they’re not always easy to get.“*

###### 2.3. Fears of inappropriate content and its impacts

Participants feared being exposed to sensitive or inappropriate images. According to P6M, the presence of content deemed inappropriate may discourage them from using these applications: *“Yes, the presence of naked people in these mobile health apps may discourage me from using them.“* P7F spoke of a disinterest in being exposed to this type of image: *“I wouldn’t like to come across images of naked women in the mobile health app.“* These fears also relate to perceived effects on psychological well-being. P12M explained: *“For example, if the mobile health app shows people who are undressed, who are French kissing, that could be bad for my mental health and that of my classmates.“* P19M mentions the negative impact of pornographic content: *“Some people put mobile apps on their phones with porn videos. That can eat at your soul.“*

###### 2.4. Perceived risks related to confidentiality and cybercrime

While weighing these risks against the perceived benefits, young people mentioned risks related to confidentiality and cybercrime, which they perceive as likely to limit their intention to use these applications. P18F stated: *“Even if these apps are beneficial, I’m a bit worried. Would it be possible for someone to use the mobile health app to locate the person and take their information?“* P19M underscored a feeling of insecurity related to the local context and the potential for disclosure of personal data: *“Yeah, we’re afraid. Because here in the Democratic Republic of Congo, nothing is secure. […] If someone sees this personal information, they may talk about it. It can be published. That can do you a lot of harm.”*

Overall, the perceived barriers to using digital CSE are very mixed. Various priorities emerge nonetheless, depending on gender: girls are more likely to mention concerns about content, confidentiality and social norms, while boys highlight technical constraints and a digital insecurity that is more general in nature. Faced with these constraints, young people develop adaptive strategies to maintain their access to digital CSE.

##### Theme 3: Financial and practical strategies for accessing digital CSE

Despite the obstacles identified in Theme 2, most of the young people expressed a clear intention to use digital CSE. This leads them to adopt concrete and ingenious adaptive strategies to circumvent the economic barriers and gain access. Such strategies can be broken down into four sub-themes.

###### 3.1. Revenue-generating activities to finance access to digital CSE

The young people described various revenue-generating activities they engage in to finance purchases of the mobile data they need to access digital CSE. P1M explained: *“This game helps me earn money. For example, I bet money on this sport, and from 1XBET, I received a good amount that allows me to pay for my megabytes.“* P8F says: *“In fact, I’m trying to make money. I sell tomatoes outside school, which generates income for me, and I’ve saved up at least 50,000 Congolese francs.”* P12M mentioned occasional or regular activities that are compatible with his schooling: *“I’m a Christian musician. I do this to earn money that allows me to pay for mobile data. And it isn’t all day long, so it has no negative impact on my schooling.”* P13M added: *“I work as a fitter-welder with my Dad during the holidays. With this job, I can get all the megabytes I need to browse the Internet and learn about sexual health.“* Some participants also mentioned personal initiatives to meet this need. P18F said: *“I also do well with my baking. This money allows me to buy data to learn about my sexual health, because I lost my mother.“* Lastly, P19M said: *“I make sure to do some small things, like selling data at my kiosk […], and I use this to pay for my own data to learn about my body.“*

###### 3.2. Cutting back on other types of spending to afford digital CSE

The young people described financial trade-offs they made that involve cutting back on certain daily expenses to prioritize buying the mobile data they need to access digital CSE. P5F said: *“Sometimes, when Mom sends me to the market, I keep the little money I have left over after shopping to pay for data. This helps me chat and get some information on menstrual cycles.“* P16M mentioned choices made to save on his transportation expenses: *“For example, when I go to school, Dad gives me money for the bus, but I try to walk to save it so that I can buy data. Yes, that’s right.”* Lastly, P21F spoke about reallocating the financial resources provided by her parents: *“My parents often give me money to pay for data so I can do my homework, and I take advantage of that to get information on how my body works.“*

###### 3.3. Use of alternative solutions to access digital CSE

Participants described using alternative solutions to access digital CSE to cope with the financial and technical constraints. These practices include using free Wi-Fi connections, portable charging devices and community charging systems. P1M stated: *“I don’t see anything that could get in the way of accessing free Wi-Fi from my neighbours.“* P4F also said: *“Sometimes I take advantage of free Wi-Fi from my neighbours.“* P12M discussed strategies to compensate for the unstable power supply: *“To make sure my phone doesn’t run out of power, I bought a Power Bank (external battery). I can use it when the power goes off so I don’t miss out on useful information about my sexual health.“* Lastly, P16M mentioned the use of community charging facilities: *“I can easily charge my phone using Kigroupe’s power supply in the neighbourhood, which costs 200 CF per charge.“*

###### 3.4. Desire for financial independence to access digital sexual and reproductive health

Congolese young people describe a strong desire for financial independence so that they can access information about their sexual health without depending on family or friends. This determination is reflected in their desire to cover the costs of accessing digital CSE on their own. P17F clearly expressed this position: *“[…] I don’t rely too much on my parents or financial assistance. I rely on myself. Yes, I take on projects. I’m interested in business, so I want to be financially independent when it comes to my sexual health. I don’t like asking for money for my Kotex (menstrual hygiene pads) every time.“* P18F mentioned some personal initiatives, however modest, to finance her purchases of mobile data: *“My small online clothing business doesn’t bring in much, but I use it to pay for my own data so I can chat and get information on my body and sexual health.“*

This theme shows that despite their young age and status as students, the participants were deploying various adaptive strategies to access digital CSE. While some of these strategies were shared by both girls and boys, there are also some distinctions: the girls tended to favour small-scale commercial activities and everyday financial trade-offs, while the boys were using a wider range of strategies, including technical or physical activities and community-based solutions.

##### Theme 4: Family and peer influences that promote the intention to use mobile health applications for CSE

This theme explores positive social influences on using digital CSE. Through three sub-themes, it presents forms of interaction and support that young people either perceive or receive from family and friends.

###### 4.1. Parental acceptance that is implicit due to cultural taboos

Participants described implicit parental acceptance of their use of mobile health applications for CSE, related to the cultural taboos around discussions of sexuality. This support is expressed less through open dialogue than through tacit approval, allowing young people to access sexual health information without violating sociocultural norms. As P7F explained: *“My parents know it’s good for me, because they wouldn’t be comfortable discussing sexuality with me in a direct way. My father wouldn’t want to talk about it directly because that’s a taboo in our culture… Yes, they would understand, because they know I can find useful information about sex that they wouldn’t want to talk about with me directly.“* Similarly, P11F emphasized this tacit approval: *“I know that Dad likes these mobile health apps because he’s embarrassed to talk to me about sex-related things and young people’s sexuality.“*

###### 4.2. Financial and technical support from family members for accessing digital CSE

In addition to the implicit approval received from parents, some family members provide financial and technical support to facilitate young people’s access to digital CSE. This support takes the form of helping install apps, covering mobile data costs, and encouraging participation in educational activities. P5F stated: *“My mother installed WhatsApp for me so that I can regularly look for information on my body’s development.“* P7F described parental support that was conditional on understanding the educational use of applications: *“My parents give me money for data, but I have to explain to them that it’s so that I can look for information about my sexual health… In fact, they let me attend a conference at Alfajiri College on November 1, 2024 on sex education for young people provided through social media. They gave me money for the bus and for data to show them that the conference was about issues related to young people’s sexuality.“* Lastly, P15F underscored the role of support from the women in her family: *“I live only with my mother and my older sister. They support me using apps to learn about my sexual health. They buy me the data packages.“*

###### 4.3. Positive peer influences on intention to use and social acceptance of digital CSE

The participants described a positive peer influence that helps strengthen their intention to use mobile health applications for CSE and social acceptance in their environment. This support is expressed through encouragement, a favourable perception of the applications and, in some cases, practical help in accessing sexual health information. P6M explained: *“Some of my classmates support me in my use of my Android phone to learn about how my genitals work, especially things that teachers don’t talk about in class because they’re embarrassed. The teachers themselves tell us to go to the Internet for information about our sexuality.“* P8F highlighted a caring and encouraging social climate: *“In my opinion, my female and male classmates have a positive perception of these mobile health apps. They encourage me to use them, they don’t make fun of me, and some of them also use the apps for sexual health information.“* This positive perception is also observed in digital exchanges between peers. P12M explained: *“In my opinion, I think they appreciate it; that’s what I’ve noticed during discussions in our WhatsApp groups.“* P13M added: *“Yes, I think my friends may also influence me to make better use of the mobile health app for my sex education. All my friends in our WhatsApp group see it as a good thing.”* Lastly, P15F spoke of the essential role played by intimate partners in this social support: *“I have a boyfriend. He cooperates so that I can use the mobile app to learn about my sexual health*.“

Overall, this theme highlights different forms of social support. Girls mainly benefit from direct family, financial, technical and relational support, provided by mothers and sisters. It facilitates the installation of applications, purchases of mobile data and access to educational spaces. Boys rely mainly on peer support in the form of encouragement, social validation and their discussions in digital groups; this promotes social acceptance and the perceived usefulness of the apps.

##### Theme 5: Perceived educational usefulness of mobile health applications for CSE

This theme, divided into four sub-themes, highlights the anticipated or proven educational benefits that young people associate with their intention to use digital CSE.

###### 5.1. Prevention of HIV and other STBBIs, adolescent pregnancy and sexual violence

Congolese youth express high educational expectations for CSE from mobile health applications. They perceive them as reliable tools for preventing risks to their sexual and reproductive health (SRH). These applications are seen as helpful for anticipating risky situations, adopting protective behaviours and reducing certain vulnerabilities. P2M explained: *“They can also help me understand how to behave with a girl to avoid getting her pregnant. And with reliable information, I can learn more about how to avoid immoral behaviour.“* P8F highlighted infection prevention: *“These mobile health applications will be very useful for protecting me against HIV and STBBIs.”* P9F emphasized the value of broader preventive information: *“They’ll give me information on the precautions to take for my sexual health […] benefits such as combatting violence, unwanted pregnancies and HIV.“* P10F mentioned the prevention of teenage pregnancies and situations of social vulnerability: *“They can give me advice on how to prevent infections and life-threatening situations such as adolescent pregnancy or unprepared marriages.“* Lastly, P12M underscored the deterrent effect of information: *“For example, in these apps, I can find information to convince me not to play around with sex, because it leads to infectious diseases.“*

###### 5.2. Access to sex education in the context of humanitarian crises and breakdowns in trust between young people and healthcare workers

For some participants, the perceived usefulness of mobile health applications for CSE increases in times of humanitarian crises and where there is a breakdown in trust with healthcare workers. In situations marked by insecurity and armed conflict, these applications are seen as an independent, accessible and discreet source of sexual health information. P8F explained: *“There’s a lot of insecurity in our country. These apps can help us girls learn about our menstrual cycles, especially when there are no doctors or midwives available due to the war in our region.“* P9F mentioned forced displacements and war as factors reinforcing the use of these digital tools: *“They’re for looking for information, especially about my sexuality in times of war. My parents live in Goma, that’s where I’m from. I left because of the rebel wars.“* P17F also spoke of a reluctance to consult health professionals, linked to discomfort in interpersonal relations: *“Doctors look at us in ways I don’t understand when we tell them about our sexual health problems. Um… I prefer to look it up on my Android phone when I’m connected.“* Lastly, P18F reported an interaction with a healthcare professional that she found intrusive and that made her anxious, reinforcing her preference for digital information, which she perceives as more discreet: *“When I lost my mother, after the funeral I went three months without a period. I went to a hospital to see a healthcare professional […]. When I explained my health problem, she started by asking me if I was pregnant. That scared me. I didn’t want to continue the conversation. […] I think a mobile app for sex education would have made it easier for me to access information without stressing me out.“*

###### 5.3. An alternative to sex education that is poor or lacking at home and at school

Several participants said they considered mobile health apps to be a direct alternative to sex education, which they report is poor or lacking at home and at school. These applications are perceived as tools that fill an educational gap by offering confidential and independent access to sexual health information. P9F explained: *“These apps can help me learn about my menstrual cycle and emergency contraception for unwanted pregnancies, topics I would never discuss with my parents or my teachers at school.“* P17F underscored the lack of spaces for discussion and the feeling of embarrassment associated with sexuality: *“I don’t get any advice on sexual health at school or from friends, for many reasons. In this mobile app, I see information that I can’t find at school or in my family. Plus, I’m ashamed to talk to anyone about sexuality right now. I just can’t do it. But it’s easy with the mobile health app.”* Lastly, P21F spoke of the role played by these apps in accessing information that parents are reluctant to share: *“We need to know things about sex, because our parents are afraid to tell us. This mobile app will help us learn about the things Mom and Dad are ashamed to tell us.“*

###### 5.4. Sharing sex education information with peers and its impact on interpersonal relationships and emotional readiness

The young people described mobile health applications as both individual learning tools and media that foster the sharing of reliable sexual health information with their peers. These applications are perceived as a means for disseminating validated content, while strengthening their discussions and interpersonal relationships. P2M explained: *“I could confirm my answers with the mobile health app before sharing them, to be sure of what I’m talking about… It also helps me inform others who don’t have access to this information.“* P3F emphasized the role played by these tools in preparing for emotional and marital life: *“I’d like my school to support the use of these apps and help educate other girls about their benefits. There are apps with advice on married life. I’m interested in that, to better manage my married life later on.“* Lastly, P19M underscored the importance of this information for social relationships and the future: *“Sex education information is important for building good relationships with my friends and for our future.“*

The perceived educational usefulness of digital CSE is therefore consistent with the personal needs expressed in Theme 1, and it is reinforced by the social dynamics described in Theme 4. The girls associated this usefulness with content related to bodily experiences, with preventing pregnancy and HIV transmission, and with managing situations of vulnerability, particularly in the context of humanitarian crises or socio-familial silence. The boys, on the other hand, placed greater emphasis on access to general prevention information, its role in raising awareness of risks, and its usefulness for sharing sexual health information in their peer groups.

##### Theme 6: Conditions for digital engagement: experience, perceived ease, accessibility

This theme presents, through three sub-themes, conditions as perceived by young people for their digital engagement, focusing on their previous experience with digital platforms and mobile applications and on the factors facilitating this projected engagement.

###### 6.1. Previous experience with digital platforms or mobile applications and their use in searching for sexual health information

Several young people described previous and regular use of digital platforms and mobile applications—such as WhatsApp, Facebook, TikTok, Instagram, Twitter, and Snapchat—both for communicating and for searching for sexual health information. This daily familiarity with digital tools is perceived as facilitating the adoption and acceptance of mobile health applications dedicated to CSE. The integrated use of mobile applications (WhatsApp) and digital platforms (Facebook) therefore appears to be a natural gateway to using digital health technologies. P10F explained: *“I regularly use WhatsApp to get news of my friends and distant family members, and I often discuss sexuality issues with my classmates in WhatsApp groups. It’s easy to download and install. If the sexual health mobile app you’re talking about is like that, then it’ll be good for us to use.“* P17F mentioned the applications she has already tried and how easy they are to access and use: *“The sexual health app I had to use was easy to download. What’s more, it was free. All I needed to be able to open it was to have my phone charged.”* P18F highlights the appeal of certain mobile apps: *“WhatsApp appeals to me. With this mobile app, I can easily find information about my sexual health and well-being.“* Finally, P19M mentioned regularly using a combination of several digital tools to find the information he needed: *“I regularly use Facebook and WhatsApp to find information about my body and my sexual health.“*

###### 6.2. Perceived ease of use of mobile health applications for CSE

Participants indicated that the perceived ease of use of mobile health applications supports their intention to use them for CSE, particularly due to the simplicity of their installation, the clarity of their content, the intuitive navigation, their notifications and their visual appeal. P1M spoke of their perceived simplicity: *“I don’t see any problems, it’s quite clear and easy to use.“* P2M affirmed the ease of interacting with their content and features: *“What would be easy is to more easily answer questions about sexual health and receive notifications.“* P3F also emphasized the accessibility of the information: *“What would be easy is if I could easily download information.“* For P6M, ease of installation would be one response to the problems encountered obtaining reliable advice from family and friends: *“Sometimes my neighbours, even my family, don’t know how to give proper sexual health advice. If we had mobile health applications that were easy to install, then that’d be a good solution for us young people.”* Lastly, P15F emphasized the importance of visual appeal and updates: *“I’m thinking about app updates. An app that’s easy for me to download, that looks good. Yes, that means a lot to me.“*

###### 6.3. Accessibility of mobile health applications for CSE: offline access, free mobile data and reliable content

Participants emphasized how the accessibility of mobile health applications is an essential condition for their intention to use them for CSE. Offline access, free or low-cost mobile data, and reliable content are seen as factors facilitating sustained engagement by limiting technical and financial constraints. P15F clearly expressed this expectation: *“I want these apps to be free and usable without an Internet plan.“* P18F mentioned the importance of being able to use the application at any time, regardless of the connection: *“Yeah, it’d be easier because you can use it at any time. I would have preferred an app that’s accessible offline, so there’d be no need for an Internet plan.“* The credibility of the information is seen as key to acceptance of these applications. P19M explained: *“If the information in mobile health apps is accurate, I can easily like them and use them. But if I see that they aren’t good, I won’t want to look at them again; I might just delete them.”* Lastly, P20F mentions the availability of free data as a facilitating factor for their use: *“What could make it easier for me to use, as a bonus, is a free connection*.”

Under this sixth theme, digital engagement as perceived by the girls was based mainly on free access, offline access, easy downloads and use, and the visual appeal of the applications. Among the boys, it was based more on clarity of use and confidence in the reliability of the information. Taken together, a cross-reading of the results shows close links between the six themes structuring Congolese young people’s intention to use mobile health applications for their SCE. The perceived barriers (Theme 2) lead young people to mobilize adaptive strategies (Theme 3) and they are mitigated by support from their families and peers (Theme 4); perceived educational usefulness (Theme 5) drives intention to use (Theme 1); and perceived conditions encourage digital engagement (Theme 6). Consequently, the intention to use digital CSE is built on the practical strategies implemented by the young people, the support they receive from families and peers, and conditions of use that are considered accessible, despite persistent economic, technical and social constraints. These perceptions are widely shared, with some nuances depending on gender. Figure 1 provides a block diagram of these relationships.

**Figure 1:**
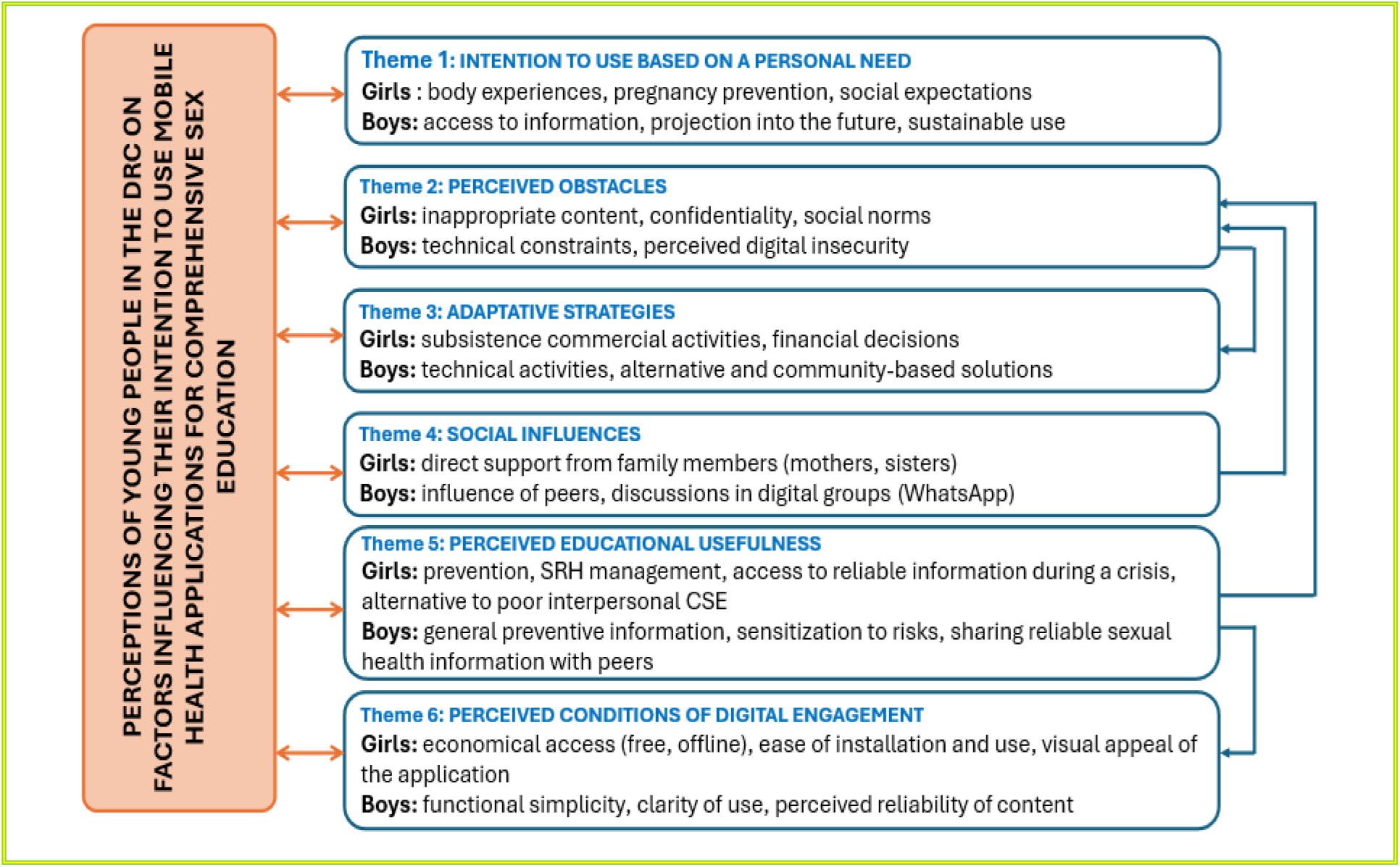
Block diagram of perceptions among young people in the DRC of the factors influencing their intention to use mobile health applications for CSE

## 4. DISCUSSION

### Key findings

Our findings show that the intention to use mobile health applications for CSE is shaped by personal motivations, material constraints, social dynamics, and contextual factors specific to Congolese society, including humanitarian crises, insecurity related to armed conflict, and disruptions of health services, effectively reinforcing the perceived value of digital solutions for maintaining educational continuity and a sense of independence.

Through the six themes identified, Congolese youth expressed their educational expectations with respect to body intelligence, HIV and STBBI prevention, contraception, rights and norms related to sexuality, combatting sexual violence, healthy interpersonal relationships, responsible decision-making and empowerment in managing their sexual health. These themes are broadly aligned with the eight components of CSE as defined in UNESCO’s revised technical guide (2). This thematic diversity suggests significant demand for sex education, persistent gaps in family and academic settings, and a clearly expressed need to fill these gaps. This need has also been recognized in an exploratory qualitative study conducted in Kenya, where young people, parents, teachers, health professionals and community volunteers perceive digital CSE as beneficial for learning, rapid access to information, knowledge improvement, educational effectiveness and narrowing the intergenerational gap (61). These findings highlight the importance of designing mobile health applications that are culturally appropriate, safe and aligned with the real needs of young people in the DRC. Other recent studies point in the same direction. A qualitative synthesis (35) shows that young people seek reliable information and are dealing with a lack of accurate knowledge on sexual health, the weight of taboos and stigma, as well as structural barriers and restrictive social norms. They attach great importance to confidentiality, which reinforces the need for appropriate and secure digital tools. A mixed systematic review (39) confirms this view: young people prefer digital tools that are simple, confidential and aligned with their specific needs. Another systematic review (36) supports the importance of varied, engaging and culturally appropriate content. Taken together, these results demonstrate that, in a context where access to institutional CSE remains limited or inadequate, mobile health applications are a promising response to young people’s educational expectations. They appear to be a space for appropriating knowledge that is often invisible in traditional educational settings.

These dynamics are clearly evident in the first theme, where the intention of young people in Bukavu to use mobile health applications is based primarily on a personal need for reliable information. Young people want to better understand their bodies, prevent risks related to their sexuality and make informed choices. Their motivation is tied to decision-making autonomy and a desire to protect themselves. This informational autonomy is part of a specific relational framework, marked by family taboos, social norms and, for some, a relationship between healthcare professionals and young people that is perceived as not safe. In this context, digital CSE becomes a relational alternative that allows for more discreet and controlled access to information. These results are consistent with those of Laar et al. (35), which show that in rural areas, young people use WhatsApp to discreetly access information on contraception, HIV and STBBIs, strengthen their autonomy and avoid stigmatization. Thus, in a variety of contexts, these tools are an essential means for obtaining information and supporting decision-making capacity in sexuality matters. In South Africa, the use of WhatsApp illustrates this dynamic: it creates a confidential and user-friendly space that promotes learning from one’s peers and strengthens decision-making capacities in sexual health (62). The perceptions observed in this study regarding the intention to use these tools on a regular basis are consistent with a logic of empowerment supported by their perceived educational value. In the young people’s discourse, this empowerment appears to be a lever that can improve their access to CSE and support their educational journey. Our results also highlight the need for an inclusive approach involving all young people, regardless of whether they own a smartphone or have ever used a mobile health application. With this in mind, our study makes an original contribution by including participants from urban and peri-urban areas, with or without smartphones and with varying levels of experience with digital technologies. This inclusive approach makes it possible to report on how young people perceive digital CSE as a service that is better suited to the diversity of their personal situations and likely to help reduce the digital divide associated with access to technology, place of residence, socioeconomic disparities and, in some cases, gender inequalities (28). However, this intention faces various obstacles, as highlighted in the second theme.

Theme 2 shows that the technological, social and economic environment strongly influences access to digital CSE. Young people reported high mobile data costs, power outages, poor connectivity, fear of inappropriate content and privacy risks. The fear of unintentional exposure to sexualized content appears to be a major barrier to young people’s intention to use. These obstacles are similar to those documented in developed and developing countries, where cyber threats, the disclosure of personal information and the mistrust they generate limit the acceptance of digital tools. In Canada, a qualitative study based on Charmaz’s constructivist grounded theory (63) shows that the sudden appearance of sexually explicit content can cause significant discomfort, rekindle negative experiences (sexual violence, stigmatization, abortion) and reduce user confidence, even when mobile health applications are intended for educational purposes. Duclos et al. (64) emphasize how in Burkina Faso, technological insecurity and lack of confidentiality hinder the use of mobile health tools dedicated to CSE. In Kenya, young people, their parents and their teachers identified the high cost of purchasing smartphones and mobile data as a significant barrier to the provision of digital CSE. This financial constraint sometimes drives young people to resort to risky strategies, including transactional sexual relations for girls and theft for boys (61). Taken together, these findings show that access to digital CSE depends as much on perceived safety as on the reliability of the technological environment, and that low socioeconomic status represents a significant barrier. However, the third theme of this study reveals that these constraints are not definitive barriers.

To finance their access to digital CSE, Congolese young people show great ingenuity in overcoming structural and financial barriers. They engage in income-generating activities, reallocate certain expenses, use free Wi-Fi, or circumvent power outages with power banks they have purchased with their own money. This proactive approach, supported by a strong social network, distinguishes our study from those conducted in other developing countries (27,36,39,61,65), which mainly describe barriers to access without highlighting young people’s active involvement in finding solutions. In Bukavu, young people do not simply accept constraints: they get organized, anticipate problems and adopt their own strategies to maintain their access to CSE. These approaches demonstrate a strong socioeconomic agency oriented toward accessing digital CSE, as well as a strong desire for financial independence and proactive adaptations to local economic and technological constraints. They are also part of a socio-familial environment marked by strong community solidarity, which emerged in the fourth theme.

Theme 4 highlights the importance of the positive influence of families and peers in the intention to use CSE mobile health applications. Parental support, implicit but real, manifests itself in a tacit acceptance that is linked to cultural taboos and in practical support: the provision of mobile data, assistance with installing mobile app, and permission to participate in scientific sex education activities. These findings are consistent with the views of parents and teachers in Kenya, for whom digital CSE is an effective and discreet way to fill the gap in sexual health information. As modesty limits direct discussions of such subjects, these actors promote digital CSE to ensure that young people will have access to reliable content and to facilitate the acquisition of smartphones by family members and friends (61). This parental investment, whether financial or technical, reflects a desire to guarantee that young people have access to high-quality CSE. It helps reduce major barriers, such as the cost of mobile data and smartphones, thereby facilitating regular and independent use of digital tools. Furthermore, in this study, participants indicated that peers and intimate partners play an important role in their intention to use these applications, particularly through material or motivational support that encourages them to search for reliable information. These social dynamics are consistent with those already reported in Africa (39), according to which peer networks legitimize the use of digital sexual health interventions and strengthen young people’s engagement. Such social normalization mechanisms help shape the intention to use digital CSE tools, whose perceived educational usefulness is confirmed in the fifth theme.

In Theme 5, Congolese young people associate mobile health applications with the prevention of HIV, STBBIs, and adolescent pregnancy as well as the management of their emotional relationships. This perception is consistent with Aydin’s analysis (66), which suggests that applications intended for Generation Z have a multidimensional preventive nature: tracking the menstrual cycle, receiving contraception reminders, learning about safe sex practices, and adopting responsible sexual health behaviours. This educational function helps reduce sexual and behavioural health risks, making young people a particularly suitable priority target for this type of tool. The perceived educational value of these applications also extends to situations of humanitarian crisis and armed conflict, where access to sexual and reproductive health services is extremely limited. They are then seen as credible alternatives to services that have been interrupted or are considered unwelcoming. These results must be interpreted in light of the occupation, by armed groups, of the cities of Goma and Bukavu at the time the data collection for this study, a situation that may have disrupted the provision of interpersonal sexual and reproductive health services (67). They are aligned with the Elrha report (68), which supports the need to deploy CSE applications in humanitarian emergencies to improve service access, quality and continuity. They also echo the perceptions of parents, health professionals and young people in Kenya, who recognize the importance of digital CSE in times of crisis: mothers value a tool that is already a part of young people’s daily lives; healthcare providers emphasize access to reliable information on STBBIs and HIV; boys see it as a means for calling for help in an emergency; and girls mention its role in socialization (61). However, the results of this study qualify these findings by revealing different perspectives according to gender. Among Congolese girls, digital CSE is perceived as a credible alternative when access to healthcare providers is limited or perceived as unsafe, a perception that is supported by implicit parental acceptance and direct support from mothers and sisters. The boys see their commitment to digital CSE as more based on the content’s perceived reliability and its usefulness in accessing sexual health information they consider relevant. Among both girls and boys, digital CSE is perceived as a means of socialization and for sharing among peers, effectively strengthening relationships, especially in times of humanitarian crisis. This relational dimension is consistent with the work of Chory et al. (69), which shows that WhatsApp groups create a sense of community, as well as with that of Jeminiwa et al. (34), which highlights young people’s preference for applications that incorporate mutual peer support. This role structuring social relationships helps maintain CSE despite disruptions in access to formal services and sheds light on the conditions that support young people’s digital engagement. These results therefore highlight the importance of considering differences between boys and girls in their experiences, support and expectations when analyzing and designing digital CSE interventions.

Theme 6 describes the perceived facilitators of digital engagement, which is based on a combination of technical and personal factors. Congolese young people are comfortable with digital CSE due to their daily use of mobile applications such as WhatsApp. While girls prefer applications that are free, accessible offline, easy to download and use, and visually appealing, boys are more interested in clarity of use and reliable content. These different preferences converge on a common expectation that digital tools should be simple, interactive and credible. The emphasis on reliability should be understood in the African context, which is marked by a high prevalence of sexual and gender-based violence facilitated by technology. A recent study conducted in seven African countries (Ethiopia, Ghana, Kenya, Liberia, Mali, Sierra Leone and Uganda) shows that 63% of girls and young women have been exposed to various forms of online violence on WhatsApp and Facebook, such as aggressive harassment, hateful content, identity abuse and cyberbullying (70). In such a risky digital environment, the perception that content is safe, reliable and relevant appears to be crucial to young people’s engagement with CSE applications. This requirement for reliability can be seen in the importance placed on clarity, personalization and interactive content, which are perceived as guarantees of credibility and information security. These factors reinforce young people’s engagement and inform their intention to use these applications on a regular basis to meet their personal information needs (34). Similar observations emerge in Kenya, where WhatsApp and Facebook are widely used by young people for communication and to search for sexual health information, due to their ease of use, the quick access they provide to content and the relevant language used (61). Such digital familiarity provides a natural gateway to acceptance of CSE-dedicated applications. In Uganda, the You Must Know application confirms this trend: young people value its ease of use and the quick access it provides to CSE services (71). Another Ugandan study shows a high usage rate (81%) that is attributed to highly appreciated technical features: non-intrusive advertising (75%), ease of use (77%), and easy-to-follow instructions (86%) (72).

Lastly, this study has strengths and limitations. Its main strength lies in its social relevance and multi-site scope. To our knowledge, it is one of the few qualitative studies on the intention to use mobile health applications for CSE by young people in FSSA, such that it provides an original contribution to this emerging field. The study’s main limitation concerns the risk of social desirability bias, mitigated by the confidentiality of the interviews and their secure setting. However, this limitation may have moderated some of the perceptions expressed on sensitive subjects. Future studies could further explore these findings in other urban or rural contexts by evaluating application prototypes in conditions of real-world use.

### Implications for research, practice and public policy

The results show young people’s ingenuity in circumventing structural barriers and their clear interest in digital CSE. For research, they highlight the need to explore intention to use in resource-limited contexts while reassessing the relevance and limitations of UTAUT. Several of the dimensions observed—expected benefits, perceived ease of use, influence of family and friends, and resource availability—are consistent with the components of the model used to formulate the interview questions. However, the study also highlights major contextual factors, such as the insecurity associated with armed conflict, humanitarian crises, and disruptions to health services, which are not part of the model. These results suggest that UTAUT should be adapted to local sociopolitical realities by incorporating structural dimensions likely to influence the acceptance of health education technologies. They also suggest that, beyond structural factors, consideration should be given to the adaptive strategies implemented by young people to cope with these constraints, which actively help shape their intention to use and their engagement with digital CSE. These results underscore the need to develop culturally anchored digital interventions that will support young people’s decision-making autonomy and protect them, as well as integrate differences in their perceptions by gender to enhance the interventions’ relevance and acceptability. For education and health policies, these results suggest the need for stronger partnerships between schools, health services and researchers to build a sustainable and equitable digital ecosystem for CSE in FSSA in general, and in the DRC in particular.

## Conclusion

This study reveals a clear intention among Congolese young people to use mobile health applications for CSE. This intention is based on the perceived educational value of digital tools, their preventive potential and their ability to provide a discreet and accessible learning space in a context where institutional CSE remains limited. It is reinforced by a supportive social environment—family members, peers and partners—that facilitates acceptance by mitigating certain technical, personal and financial barriers. Young people also demonstrate a remarkable ability to adapt to economic and technological constraints, particularly through alternative access strategies. This ingenuity, rarely documented in low-resource countries, reveals their active role in building an inclusive digital environment for CSE in the DRC.

The major contribution of this study lies in the in-depth understanding it has developed of perceptions about the factors influencing intention to use in a context of precariousness, an area that is relatively unexplored in CSE. These findings can inform the design of relevant, acceptable and culturally rooted digital interventions, in particular the development and implementation of mobile health applications that are more accessible and reliable and better tailored to local realities. Such interventions will benefit from incorporating gender differences in perceptions and expectations, as revealed by this study, to enhance their relevance, acceptability and effectiveness. By encouraging the active involvement of young people in the co-design of solutions, this study contributes to the field of digital health and behavioural interventions. Lastly, it supports incorporating digital CSE into educational and health strategies to enhance equal access and strengthen young people’s decision-making autonomy in an environment characterized by accelerated digital transformation.

## Data Availability

The data, including verbatim transcripts from individual interviews with young people in Bukavu, are not publicly available due to their sensitive nature. However, they may be obtained from the corresponding author upon reasonable request.

## Acknowledgements

We would like to express our deepest gratitude to the Provincial Inspector of the Ministry of National Education and New Citizenship of the Democratic Republic of the Congo, South Kivu Province, for the institutional support it provided throughout this study. We also thank the principals of the participating schools and the focal point teachers for their collaboration, availability, and commitment to coordinating data collection. Lastly, we extend our warmest thanks to the young students who took part in the individual interviews. Their availability, trust and enthusiasm in sharing their perceptions were essential to the completion of this study.

## Some abbreviations

COREQ: Consolidated criteria for reporting qualitative research
CSE: Comprehensive sex education
DRC: Democratic Republic of Congo
FSSA: French-speaking sub-Saharan Africa
HIV: Human Immunodeficiency Virus
SRH: Sexual and reproductive health
STBBI: Sexually transmitted and blood-borne infection
UNESCO: United Nations Educational, Scientific and Cultural Organization
UNFPA: United Nations Population Fund
UNICEF: United Nations Children’s Fund
UTAUT: Unified Theory of Acceptance and Use of Technology

## Supporting information

### S1: COREQ: 32-item checklist

Developed from: Tong A, Sainsbury P, Craig J. Consolidated criteria for reporting qualitative research (COREQ): a 32-item checklist for interviews and focus groups. International Journal for Quality in Health Care. Dec. 1, 2007;19(6):349-57.

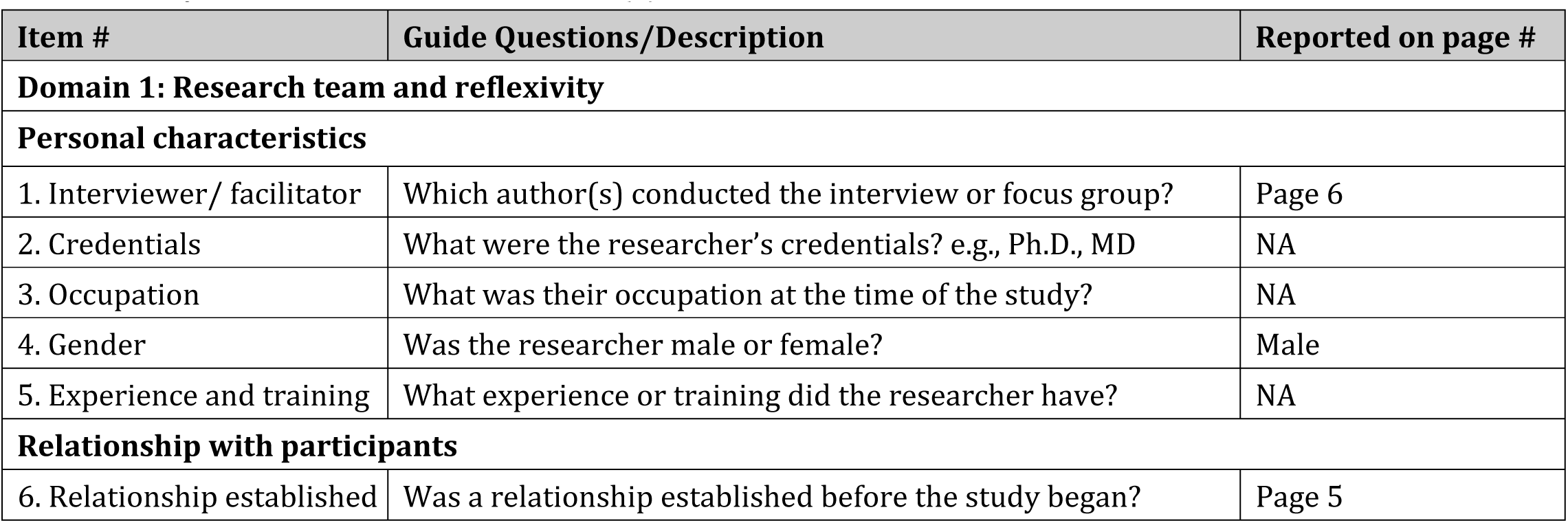
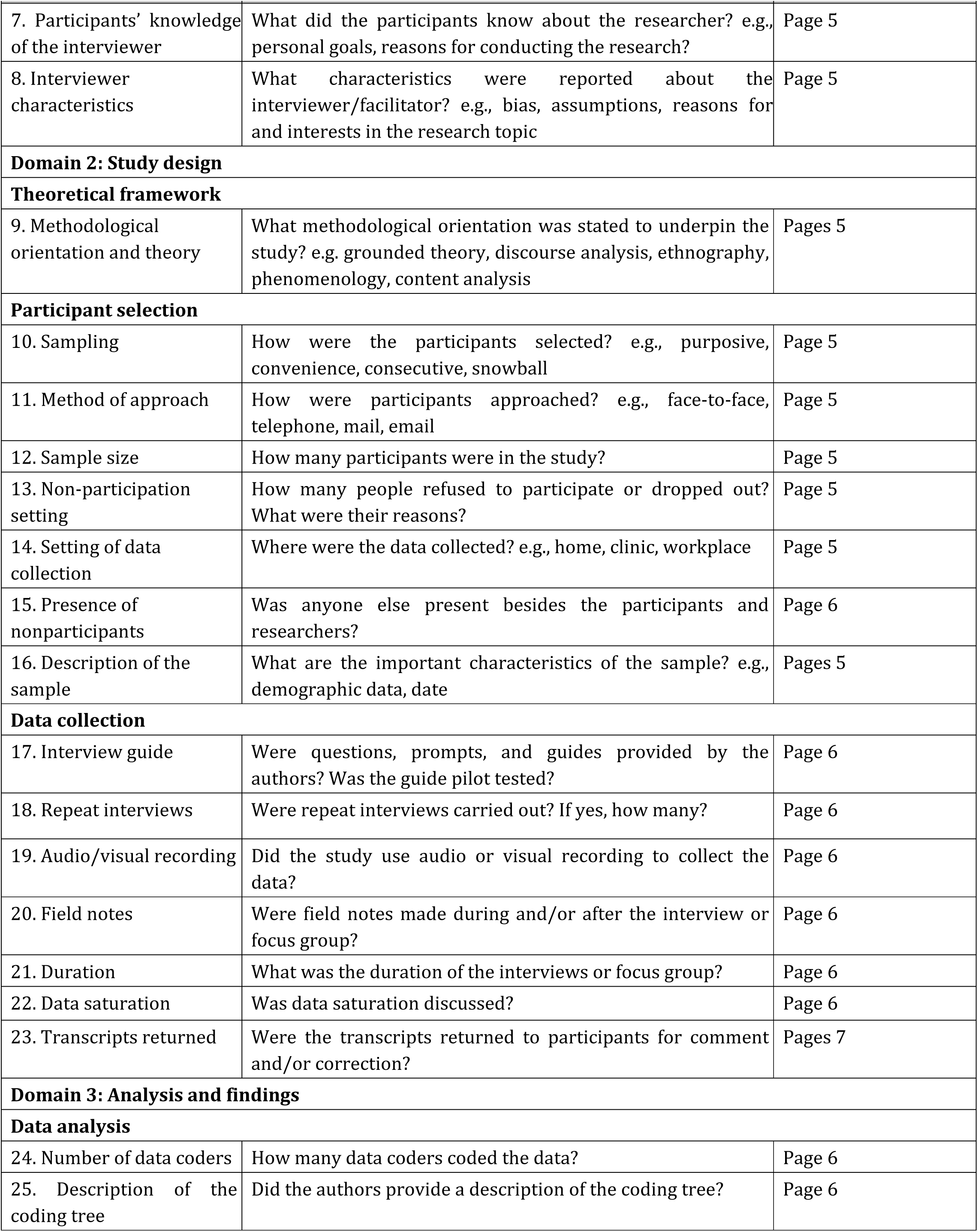
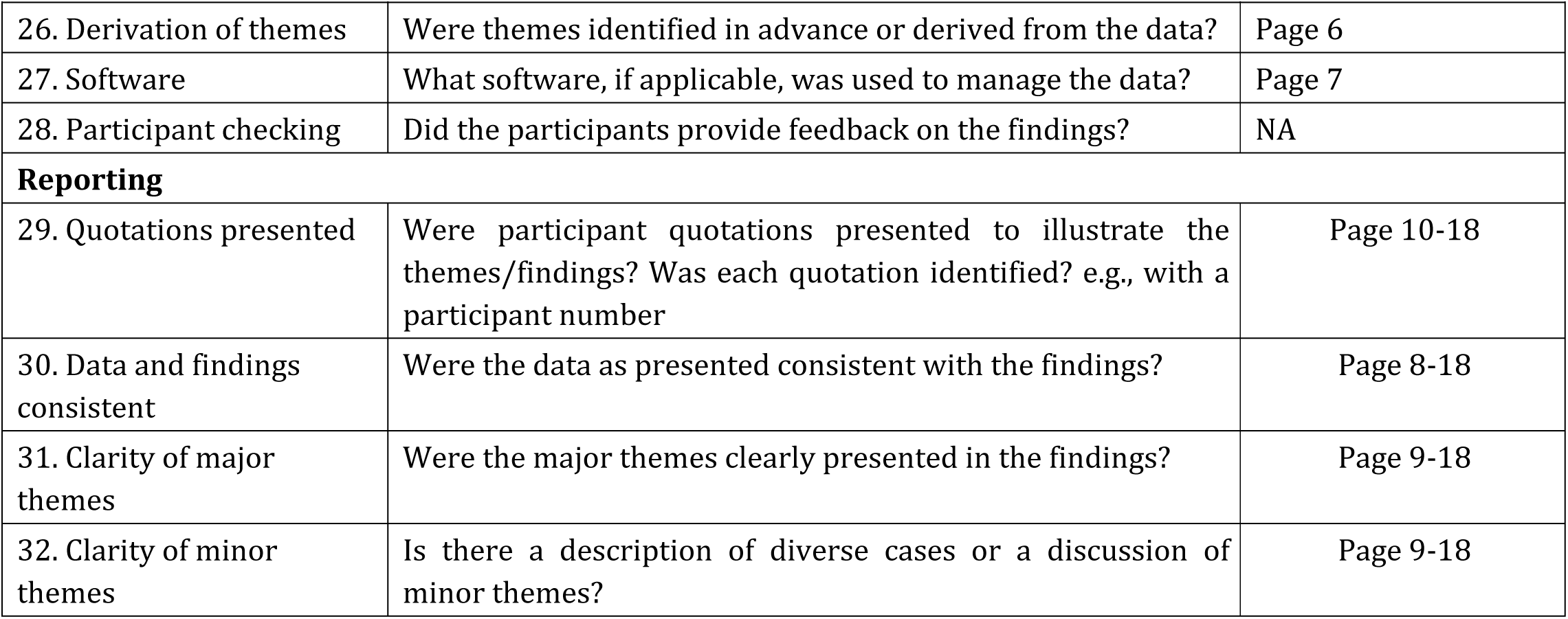

### S2: Interview guide

Perceptions of young people in the Democratic Republic of the Congo on the factors influencing their intention to use mobile health applications for comprehensive sex education: A descriptive qualitative study

#### I. Sociodemographic and educational characteristics of the participants

i. Age in years:
ii. Sex at birth:
iii. Place of residence (municipality and neighbourhood):
iv. School attended:
v. Stream:
vi. Level of education (specify class):
vii. Access to and use of mobile technologies (ownership of a smartphone or tablet).

#### II. Participants’ perceptions of factors influencing their intention to use mobile health applications for CSE

1. To what extent are you willing to use mobile health applications on a regular basis for your comprehensive sex education?
2. What factors could encourage you to use these applications on an ongoing basis?
3. What benefits could the use of mobile health applications have for your comprehensive sex education?
4. How do you think these applications could improve your knowledge of sexual health?
5. In your opinion, what would be easy about using mobile health applications? What would be difficult?
6. What resources would you have for using these applications?
7. What support could you receive from family and friends that would help you use these technologies? What could hinder the support from family and friends?
8. How do your friends and classmates feel about you using mobile health apps for your comprehensive sex education?
9. Can you explain how the influence of friends and family might affect your decision to use these apps?
10. What concerns might you have about the information you would share through these apps? If necessary, do you think about privacy or security issues?

#### III. Conclusion

Is there anything else that we haven’t discussed that you would like to add? Or what do you think is the most important thing to remember about using a mobile app for your comprehensive sex education?

Thank you for your participation!

## Notes

### Competing Interest Statement

The authors have declared no competing interest.

### Author Declarations

This study was approved by the Research Ethics Committee in Science and Health at Université de Montréal (CERSES 2024-6039) and by the National Health Ethics Committee of the Democratic Republic of Congo (CNES 001/DPSK/222PM/2024). Participation was voluntary and conducted in accordance with confidentiality requirements. Written informed consent was obtained from participants aged 18 years and older. For participants aged 15–17 years, consent was obtained through the school authorities acting as legal guardians in accordance with local regulations, and assent was obtained from the students prior to participation.

